# Design and proof-of-concept evaluation of a touchless connector system for preventing peritoneal dialysis-associated peritonitis

**DOI:** 10.1101/2021.09.20.21263563

**Authors:** Ibrahim O. Yekinni, Tom Viker, Ryan Hunter, Aaron Tucker, Sarah Elfering, Michelle N. Rheault, Arthur Erdman

## Abstract

**Introduction:** In this paper, we describe the design of a touchless peritoneal dialysis connector system and how we evaluated its potential for preventing peritoneal dialysis-associated peritonitis, in comparison to the standard of care. The unique feature of this system is an enclosure within which patients can connect and disconnect for therapy, protecting their peritoneal catheters from touch or aerosols.

**Methods:** We simulated a worst-case contamination scenario by spraying 40mL of a standardized inoculum [1×10^7^ colony-forming units (CFU) per milliliter] of test organisms, *Staphylococcus epidermidis* ATCC1228 and *Pseudomonas aeruginosa* ATCC39327, while test participants made mock connections for therapy. We then compared the incidence of fluid path contamination by test organisms in the touchless connector system and the standard of care. 4 participants were recruited to perform a total of 56 tests, divided in a 1:1 ratio between both systems. Peritoneal dialysis fluid sample from each test was collected and maintained at body temperature (37° C) for 16 hours before being plated on Luria Bertani agar, Mannitol Salts Agar and Pseudomonas isolation agar for enumeration.

**Results:** No contamination was observed in test samples from the touchless connector system, compared to 65%, 75% and 70% incidence contamination for the standard of care on Luria Bertani agar, Mannitol Salts Agar and Pseudomonas isolation agar respectively.

**Conclusion:** Results show that the touchless connector system can prevent fluid path contamination even in heavy bacterial exposures and may help reduce peritoneal dialysis-associated peritonitis risks from inadvertent contamination with further development.

SUMMARY BOX

What are the new findings?
We describe the design and evaluation of a touchless peritoneal dialysis connector system showing reduced incidence of bacterial contamination in comparison to the standard of care.

How might it impact on healthcare in the future?
Integrating the touchless connector system into peritoneal dialysis systems may help reduce peritonitis risks in patients.

## 1. BACKGROUND

### 1.1 Introduction

Peritoneal dialysis (PD) is one of the options available for treating end-stage kidney disease, the last stage of chronic kidney disease in which the organs no longer work enough for a patient to survive without dialysis or a transplant. In the option most people know, hemodialysis, patients visit a dialysis center thrice weekly to use a machine that filters excess fluid and solutes from their bloodstream. In PD, these excess fluids and solutes are filtered via a membrane in the abdomen called the peritoneum. The space of the peritoneum is filled with dialysis fluid through a catheter implanted in the abdomen (the *permanent peritoneal catheter*); osmotic and concentration gradients between the blood and the fluid lead to the removal of excess body fluids and solutes, respectively. It is the preferred option for dialysis for children with blood volumes too small for circulation in a hemodialysis machine.

There is renewed interest in PD in both developed and developing countries. In the United States for example, a new payment model introduced by the Centers for Medicare and Medicaid Services is incentivizing physicians and providers to increase the proportion of patients on home therapies like PD.[1] The COVID-19 pandemic and the desire for social distancing also increased PD demand.[2]

In developing countries, recent studies have drawn attention to how limited access to kidney replacement therapy may result in up to 7 million deaths yearly, more than the deaths from HIV and Tuberculosis combined. Less than a third of patients in Asia and only 16% of patients in Africa are said to receive needed treatment, mostly due to unaffordable costs.[3] In the 2019 Global Kidney Health Atlas, the International Society of Nephrology has recommended a peritoneal dialysis first treatment approach as a potential solution to this problem.[4] Despite the potential benefits, PD uptake continues to be limited and this is due in part to concerns about peritonitis - an infection of the peritoneal membrane through which the therapy occurs. Almost a century since Georg Ganter first described PD in a 1923 paper,[5] peritonitis continues to be a common and serious complication globally.[6,7] It is the direct or major contributing cause of death in >15% of patients and the most common reason patients stop the therapy. Peritonitis leads to repeat episodes in 3% - 20% (14% overall), surgeries for peritoneal catheter removal in 10% - 88% (22% overall), permanent transfer to hemodialysis in 9% - 74% (18% overall) and death in 0.9% - 8.6% (2% - 6% overall).[8-11] Strategies to prevent peritonitis are urgently needed.

### 1.2 Peritonitis, Causes and Prevention

There are different causes for peritonitis, including catheter contamination, infection of catheter exit site or tunnel, intestinal bacteria, bacteria in patient blood or even infections ascending from uterine or vaginal sources.[12] However, contamination of the peritoneal catheter while patients connect for treatment is the most common cause. More than 50% of peritonitis episodes are caused by bacteria colonizing the skin or mucous membranes and touch contamination alone contributes to over 40% of peritonitis episodes.[13, 14]

In a 2018 review, Salzer reported that “*Most cases of PD-related peritonitis are the result of touch contamination*” and this was consistent with findings from the 2016 report of the Standardizing Care to Improve Outcomes in Pediatric ESRD (SCOPE) where touch contamination was associated with a higher risk of peritonitis (RR, 2.22; 95% CI, 1.44 to 3.34).[15,16] Contamination of the catheter by respiratory droplets is also an important source as nasal bacterial carriage can increase risk of peritonitis in patients. When compared with noncarriers, carriers had a 2 – 6-fold higher incidence of *Staphylococcus aureus* peritonitis,[17] and studies using phage typing of *S. aureus* from the patients with peritonitis found that in most cases, the isolates were the same as in the nostrils.[18-20]

The International Society for Peritoneal Dialysis (ISPD), a global authority on PD, which has published recommendations for treatment and prevention of peritonitis for over 30 years recommends that peritonitis rates for a dialysis center be below 0.5 episodes per patient-year. This means that for a dialysis provider with 100 patients undergoing PD for a year, the total number of peritonitis episodes recorded in that year should be below 50. Yet, the rate reported from providers in different countries and within the same country continues to vary widely. For example, reported rates range from 0.06 episodes per patient-year in Taiwan to 1.66 episodes per patient-year in Israel and there have been up to 20-fold variation in the reported rates within Australia, Austria, Scotland and the United Kingdom. While the sources of these variations are poorly investigated, they may be due to factors like each provider’s patient population or their PD training practices.[11]

In the most recent guidelines,[21] the ISPD recommends rigorous training in aseptic technique to reduce rates from patient-related sources of peritonitis. Providers also train new patients and retrain them after peritonitis episodes. The patients are expected to report any breach in the aseptic technique, so further actions can be taken to prevent peritonitis e.g. changing the catheter extension sets, or administering antibiotics.[22]

This reliance on patients to control peritonitis is however problematic, as fewer than 50% of patients remain adherent to their training after six months of commencing therapy.[23] This might also explain why many providers restrict PD to patients whom they feel will comply with training while excluding many otherwise eligible patients.[24]

Essentially, the wide variation in peritonitis rates and the selective prescription of PD reveal gaps in peritonitis prevention and the need for innovation. The increasing demands for PD across the world, from payment changes in favor of cost-effective home therapies, to the attempts at increasing access and affordability of dialysis in developing countries, make it even more crucial that solutions that better prevent peritonitis be developed.

In this paper, we describe the design and proof-of-concept evaluation of a touchless connector system that has the potential to reduce PD-associated peritonitis.

## 2. METHODS

### 2.1 Premise and Design

The theory behind the design of the touchless connector system is that eliminating inadvertent contamination of the peritoneal catheter will reduce peritonitis episodes by up to 50%, the proportion of peritonitis episodes caused by skin or mucous membrane bacteria.[13]

Studies investigating how different connection points in a PD system contribute to peritonitis are limited but by interviewing PD nurses, we found the *catheter extension set* (also called *transfer set*) to be a vulnerable connection point common to both manual and automated devices. In manual PD, called continuous ambulatory peritoneal dialysis (CAPD), the *catheter extension set* is the only connection point when a *double bag* (fluid bag with pre-attached drain bag) is used. The potential for contamination may also be higher since these patients have to connect their *catheter extension set* for therapy multiple times daily. In automated PD (APD) using a cycler, patients typically connect their *catheter extension set* for therapy once daily but there are other vulnerable connection points e.g., connection to fluid bags, drainage bags, etc. However, there is evidence suggesting that the automated flush-before-fill mechanisms of the cyclers may reduce contamination,[25] leaving the *catheter extension set* as the remaining vulnerable component in this modality as well.

Based on these findings, the functional need was established as “**a way to help PD patients connect/disconnect *catheter extension sets* for therapy without contamination**”.

In existing PD systems, the *catheter extension set* extends from the *permanent catheter* implanted in the patient’s abdomen **(Figure 1)**. It has a disposable *disconnect cap* which protects it from contamination when not being used for therapy. Every day, or multiple times daily in CAPD, when patients transfer dialysis fluid into their peritoneum, they first remove and discard this *disconnect cap* before connecting their *catheter extension set* to the *dialysis fluid source*. After completing therapy, they disconnect from the *dialysis fluid source* and cover the *catheter extension set* with a new *disconnect cap*. Depending on patient compliance with training, these moments when the *catheter extension set* is open could result in contamination of the peritoneal catheter and eventual peritonitis.

**Figure 1:**
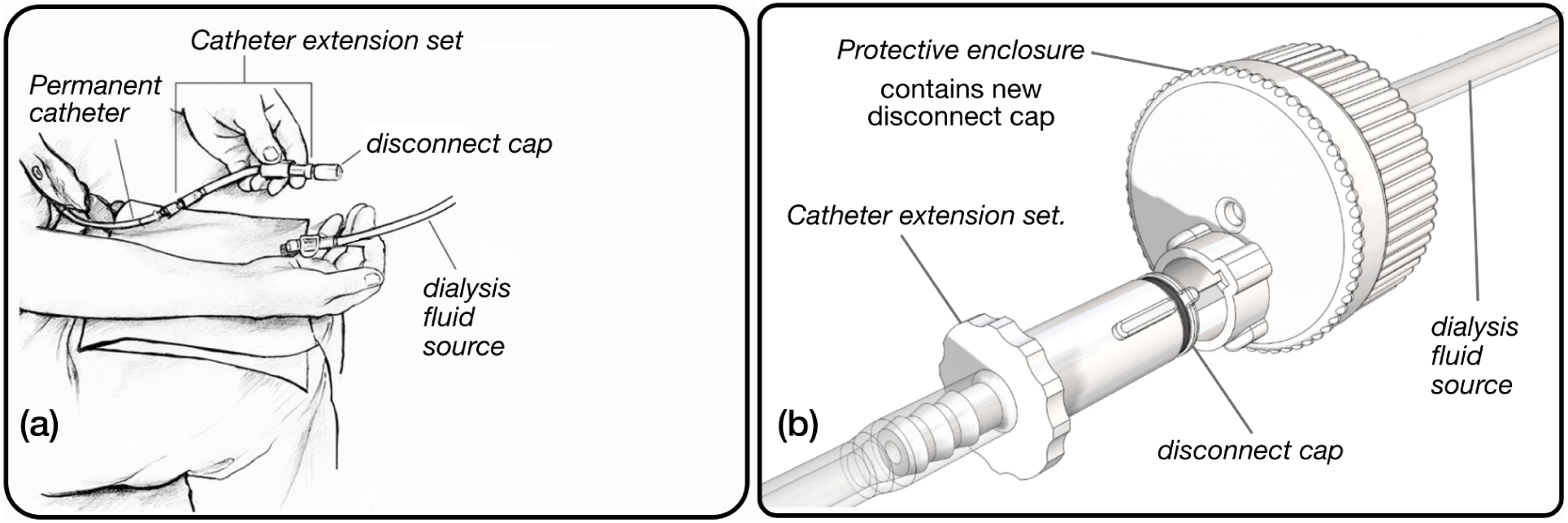
Peritoneal dialysis connector system designs (a) Standard of care connector system (b) Touchless connector system

In order to meet the established need, we identified six design criteria which drove the development of the touchless connector system.

1. Remove the old *disconnect cap* on the *catheter extension set* without physical contact
2. Eliminate contact between the old, contaminated *disconnect cap* and the *catheter extension set* after cap removal
3. Connect and disconnect the *catheter extension set* to the *dialysis fluid source* without physically contacting either
4. Connect a new, sterile *disconnect cap* to the *catheter extension set* after therapy without physically contacting either
5. Protect the entire process from aerosols via an enclosure
6. Provide patients with a step-by-step, error free process for connecting their *catheter extension set* to a *dialysis fluid source*.

Multiple ideas were generated, screened and optimized before arriving at the underlying concept for the touchless connector system. In this concept, the components of interest i.e., *catheter extension set, disconnect cap*, and *connector to the dialysis fluid source* have been redesigned as shown in **Figure 1**.

The operation of the touchless connector system **(Table 1)** is similar to that described above for existing systems. However, in this design, removal of old *disconnect caps* from the *catheter extension set*; connection of the *catheter extension set* to *dialysis fluid source* for fluid exchange; and recapping of the *catheter extension set* with a new *disconnect cap*, all take place within a *protective enclosure*. This enclosure is unique to the touchless connector system and is a redesign of the connector to “*dialysis fluid source*” in existing systems **(Figure 1)**. It also contains a new, sterile *disconnect cap* that will cover the *catheter extension set* after therapy is completed. It is a disposable component discarded with the *dialysis fluid source* (fluid bag in CAPD or cycler sets in APD) and used *disconnect cap* after therapy.

**Table 1:** Connecting and disconnecting for therapy with touchless connector system.

|  |  |
| --- | --- |
| **1. Connecting catheter for therapy** | **2. Disconnecting catheter after therapy** |
| (i) Insert *catheter extension set* into *protective enclosure* | (i) Close valve to stop flow |
| (ii) Twist the *catheter extension set* to unlock old *disconnect cap* | (ii) Pull *catheter extension set* to disconnect from *dialysis fluid source* |
| (iii) Pull the *catheter extension set* to remove old *disconnect cap* | (iii) Rotate base of *protective enclosure* |
| (iv) Rotate base of *protective enclosure* | (iv) Push in *catheter extension set* to add new, sterile *disconnect cap* |
| (v) Push in *catheter extension set* to connect with *dialysis fluid source* | (v) Twist *catheter extension set* to lock new *disconnect cap* |
| (vi) Open valve to allow flow | (vi) Withdraw capped *catheter extension set* from *protective enclosure* |

To develop the concept, we used an iterative design methodology, advancing through multiple 3D printed prototypes **(Figure 2)** based on feedback from nephrologists, nurses, patients, large dialysis organizations, and manufacturers. In total, 118 interviews were conducted across the United States between June and November 2019. The findings from 57 of these interviews with PD providers and an anonymous survey in 6 peritoneal dialysis centers helped validate the functional need and value proposition. From all interviews, the team gathered insights on required device characteristics, manufacturing goals and compatibility with existing PD systems. In fall 2020, the team conducted a usability study with 7 participants to understand and optimize usability of the touchless connector system for patients.

**Figure 2:**
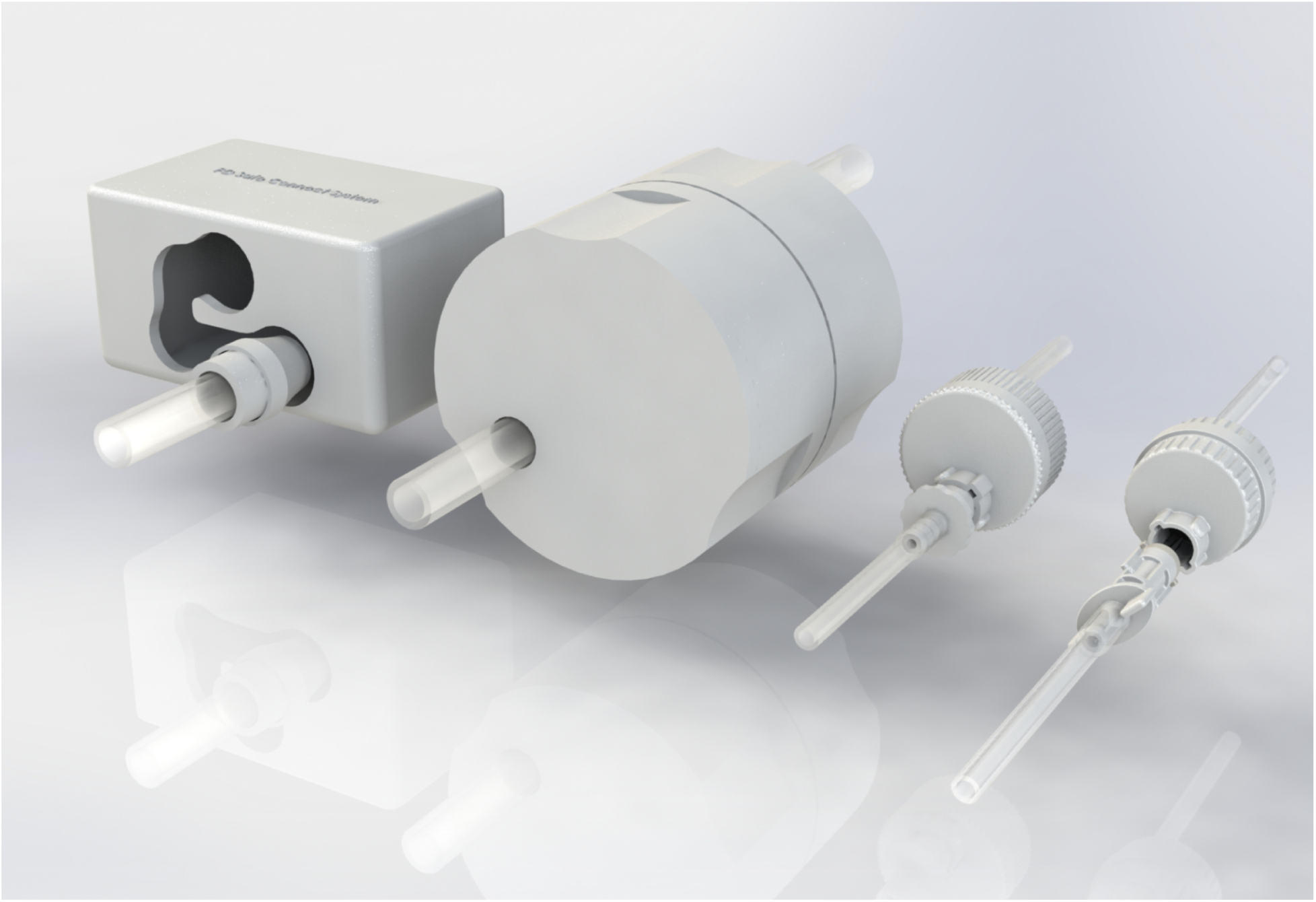
3D rendering showing design evolution of the touchless connector system

The outcome of this iterative design process is the current injection molded prototype **(Figure 3)** of the touchless connector system used in the proof-of-concept evaluation.

**Figure 3:**
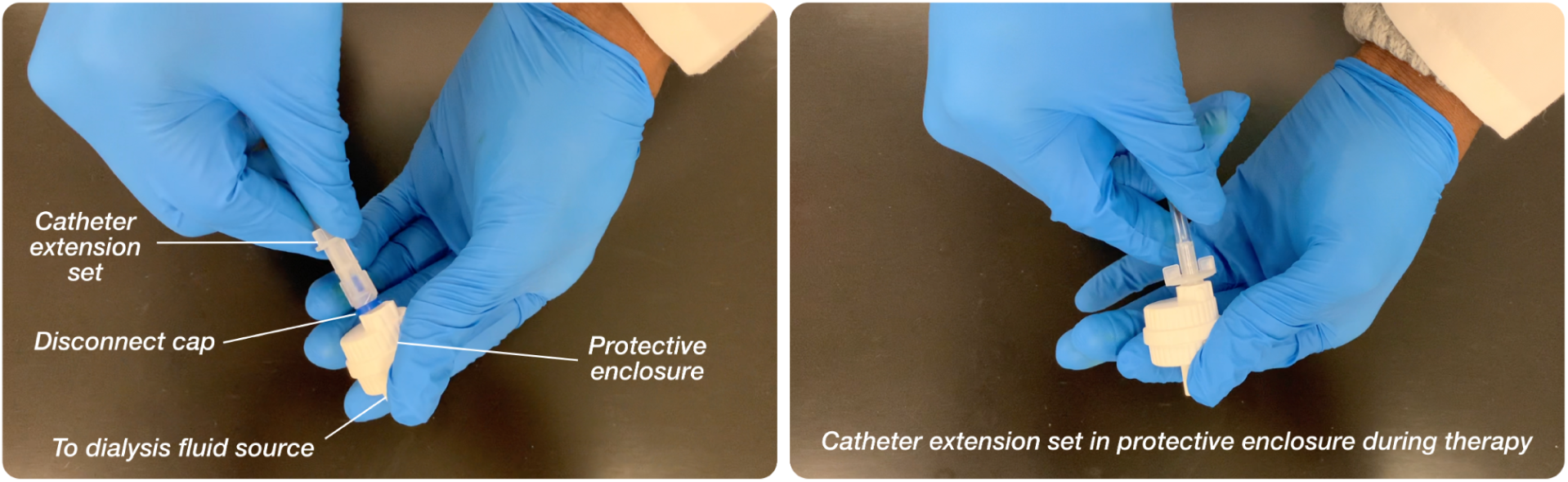
Current injection molded prototype of touchless connector system

### 2.2 Proof-of-concept Evaluation

#### 2.2.1 Rationale/Study Aims

Since the touchless connector system differs from the standard of care in its ability to prevent touch/aerosol contamination during therapy, we sought to evaluate this experimentally. A reduction of peritonitis episodes will be the most desired validation but this is not practical with current development stage and resources. We’ve therefore used an approach that simulates a worst-case scenario and assesses the ability of connector systems to prevent contamination. Appropriate ethical and biosafety clearance was obtained from the University of Minnesota Institutional Review Board and Biosafety Committee respectively (IRB ID: STUDY00010459).

#### 2.2.2 Materials

Touchless connector system prototypes; Samples of a commonly used connector system (standard of care); Sterile dialysis fluid bags; 100mL effluent sample bags. Test devices for both systems were pre-assembled and sterilized with ethylene oxide before being used to form the setup in **Figure 4**.

**Figure 4:**
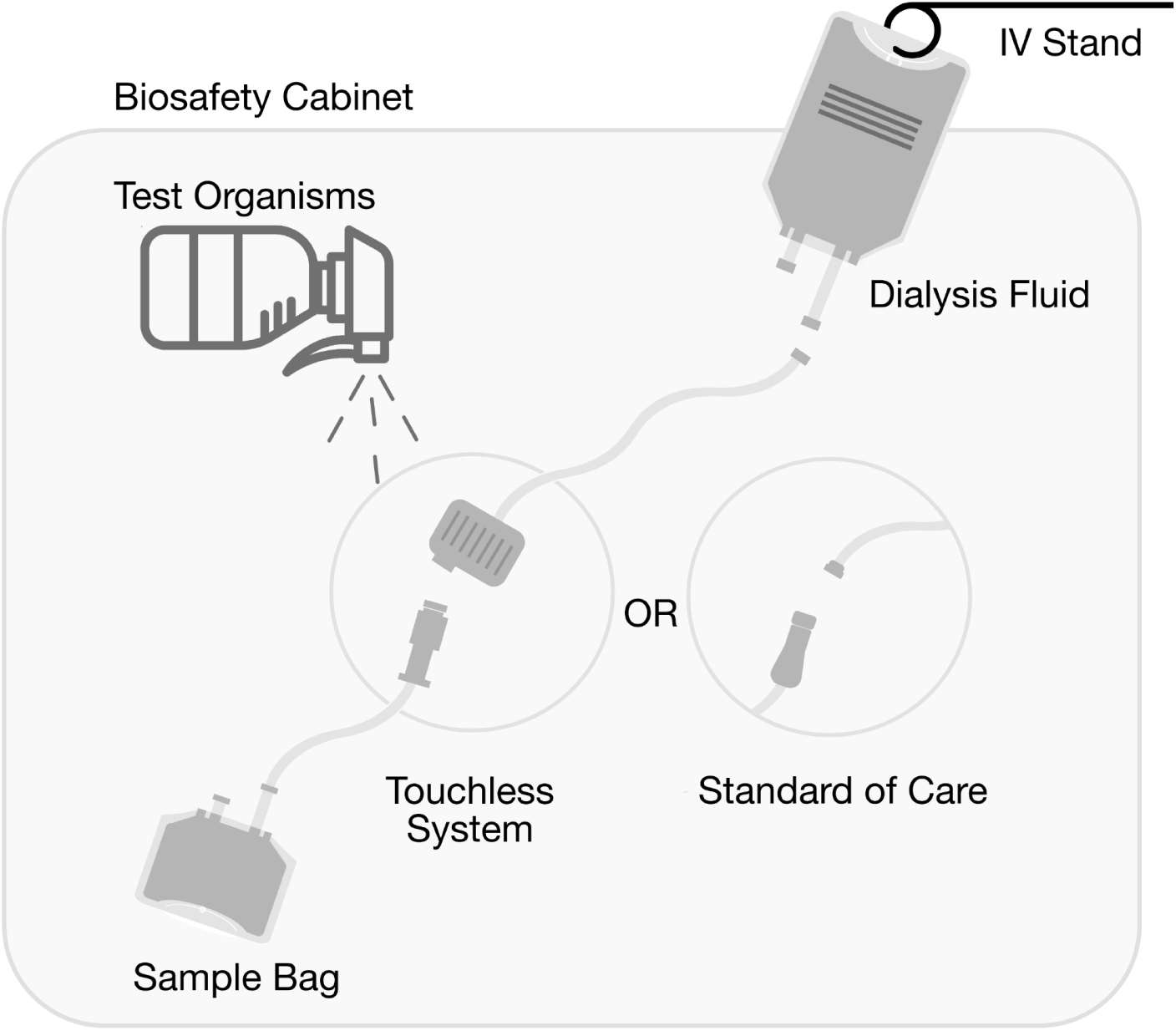
Experimental setup

*Staphylococcus epidermidis* ATCC1228 and *Pseudomonas aeruginosa* ATCC39327, representing the most commonly observed Gram-positive and Gram-negative bacteria respectively in peritonitis,[26] were used as the test organisms.

#### 2.2.3 Experimental Setup

The experiments were performed within a laminar flow biosafety cabinet in a biosafety level 2 laboratory facility to control the test environment. Experimental setup **(Figure 4)** included a dialysis fluid bag suspended from an IV pole, and connected to one end of the test devices i.e., the *dialysis fluid source* connection, making it available for participants within the biosafety cabinet. The other end of the test devices consisted of the *catheter extension set* for the touchless connector system or standard of care pre-attached to the effluent sample bags. The effluent sample bags represent the peritoneal space into which dialysis fluid will flow during therapy, and within which contaminating organisms will cause peritonitis.

To simulate contamination, 40mL of a standardized inoculum [1×10^7^ colony-forming units (CFU) per milliliter] of test organisms was sprayed while test participants made connections. To specifically assess resistance to contamination, the inoculum was sprayed when the used *disconnect caps* in both systems had been removed and the *catheter extension sets* were open. After test organisms had been sprayed, participants then allowed dialysis fluid to flow from the fluid bag, through the *catheter extension sets*, into the effluent sample bags. The sample bags were then disconnected from the system for microbiology evaluation and the process repeated for all tests. IOY and TV validated this experimental method with a prior study of 14 tests.

#### 2.2.4 Sample Size, Test Participants and Test Sequence

According to Virzi, 80% of all usability issues are detected with at least 4 participants for a minimum sample size of 4.[27] 4 non-patient adults were recruited with fully informed consents to perform the experiment. Non-patient participants were preferred because of their similarity to new PD patients. The participants received training about PD and the use of both systems.

They were allowed to practice till they felt comfortable using both systems as will be necessary for the experiment.

For each participant, test sequence for both touchless connector system and the standard of care included 7 samples obtained as follows:

i. Sample 1: a sterile control before exposure to test organisms;
ii. Samples 2 – 6: while the 40mLs of test organisms are sprayed;
iii. Sample 7: a contaminated control in which 100mL of the standardized inoculum deposited in 2 Liters of dialysis fluid is allowed to flow through the fluid path of the connection systems. To minimize bias, tests were alternated between both systems for each participant.

#### 2.2.5 Microbiology

*Pseudomonas aeruginosa* ATCC39327 (PA) and *Staphylococcus epidermidis* ATCC1228 (SE) were purchased from Microbiologix (St. Cloud, MN) and were routinely cultured and maintained on Luria Bertani (LB) agar. Strains were cultured overnight in Luria Bertani medium (6 × 5ml cultures of each bacterium).

Overnight cultures were combined and centrifuged at 7,000 × *g* for 10 minutes at 4°C. Cell pellets were washed once in 30mL of sterile phosphate buffered saline (PBS) followed by another centrifugation at 7,000 × *g* for 10 minutes. Samples were then combined (PA + SE) and resuspended in 600mL of sterile PBS to a final OD600 of 0.1, corresponding to ∼1 × 10^7^ to 1 × 10^8^ cells of each organism. Final bacterial suspensions were prepared less than 30 minutes prior to transfer to the University of Minnesota Bio-Nano Lab for the experiments. Upon delivery of sample bags after the experiments, they were immediately placed in a 37°C incubator to mimic human body temperature. The sample bags were stored in the incubator for 16 hours prior to analysis.

For the analysis, 200uL of dialysis fluid was removed from each sample and placed in a 96 well microtiter plate. Serial dilutions of each sample were performed (10^−1^ to 10^−8^) by adding 20uL of the sample into 180uL of sterile PBS. 10uL of each serial dilution were plated on Luria Bertani agar (for total bacterial enumeration), Mannitol Salts Agar (for *Staphylococcus* enumeration), and Pseudomonas isolation agar (for *Pseudomonas* enumeration). Plates were incubated at 37°C overnight, and colonies were counted the next day.

### 2.3 Patient and Public Involvement

Patients and other stakeholders were involved in the iterative design of the touchless connector system but were not involved in the design of the proof-of-concept evaluation.

## 3. RESULTS

Participants were between ages 30 and 60, with training time ranging from 5 mins to 13 minutes **(Table 2)**. None had previous PD experience and they all completed required tests for both systems.

**Table 2:**
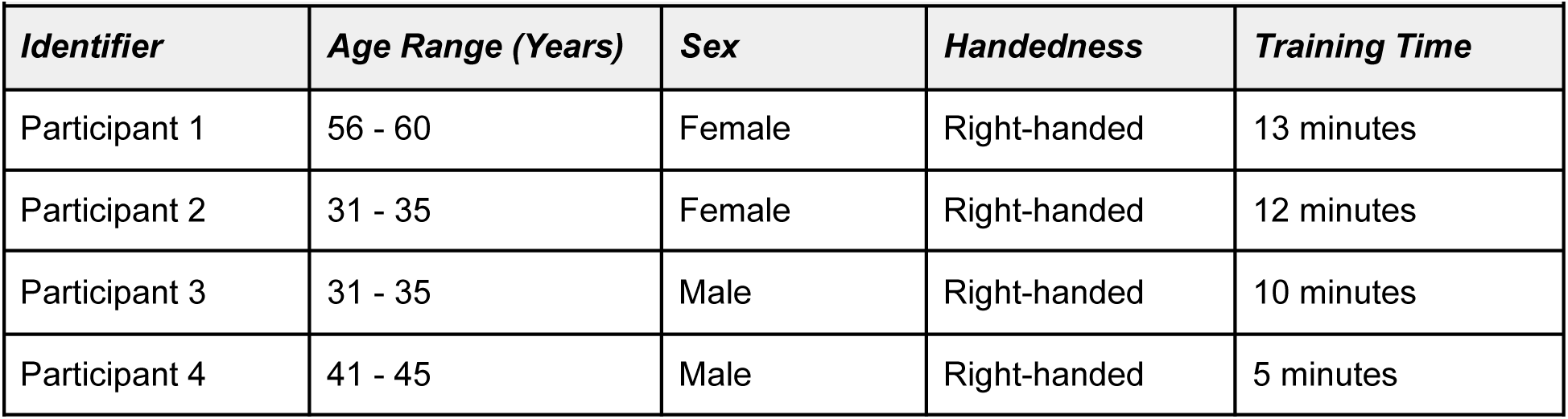
Participants Characteristics and Training Time.

| **Table 2: Participants Characteristics and Training Time** |  |  |  |  |
| --- | --- | --- | --- | --- |
| *Identifier* | *Age Range (Years)* | *Sex* | *Handedness* | *Training Time* |
| Participant 1 | 56 - 60 | Female | Right-handed | 13 minutes |
| Participant 2 | 31 - 35 | Female | Right-handed | 12 minutes |
| Participant 3 | 31 - 35 | Male | Right-handed | 10 minutes |
| Participant 4 | 41 - 45 | Male | Right-handed | 5 minutes |

For the microbiology test results, sterile and contaminated controls (Samples 1 and 7) for both systems returned expected results, i.e. observed bacterial growths on plates cultured from contaminated controls (Sample 7) and no observable growths on plates from sterile controls (Sample 1). Enumeration for remaining tests is as shown in **Table 3**.

**Table 3:**
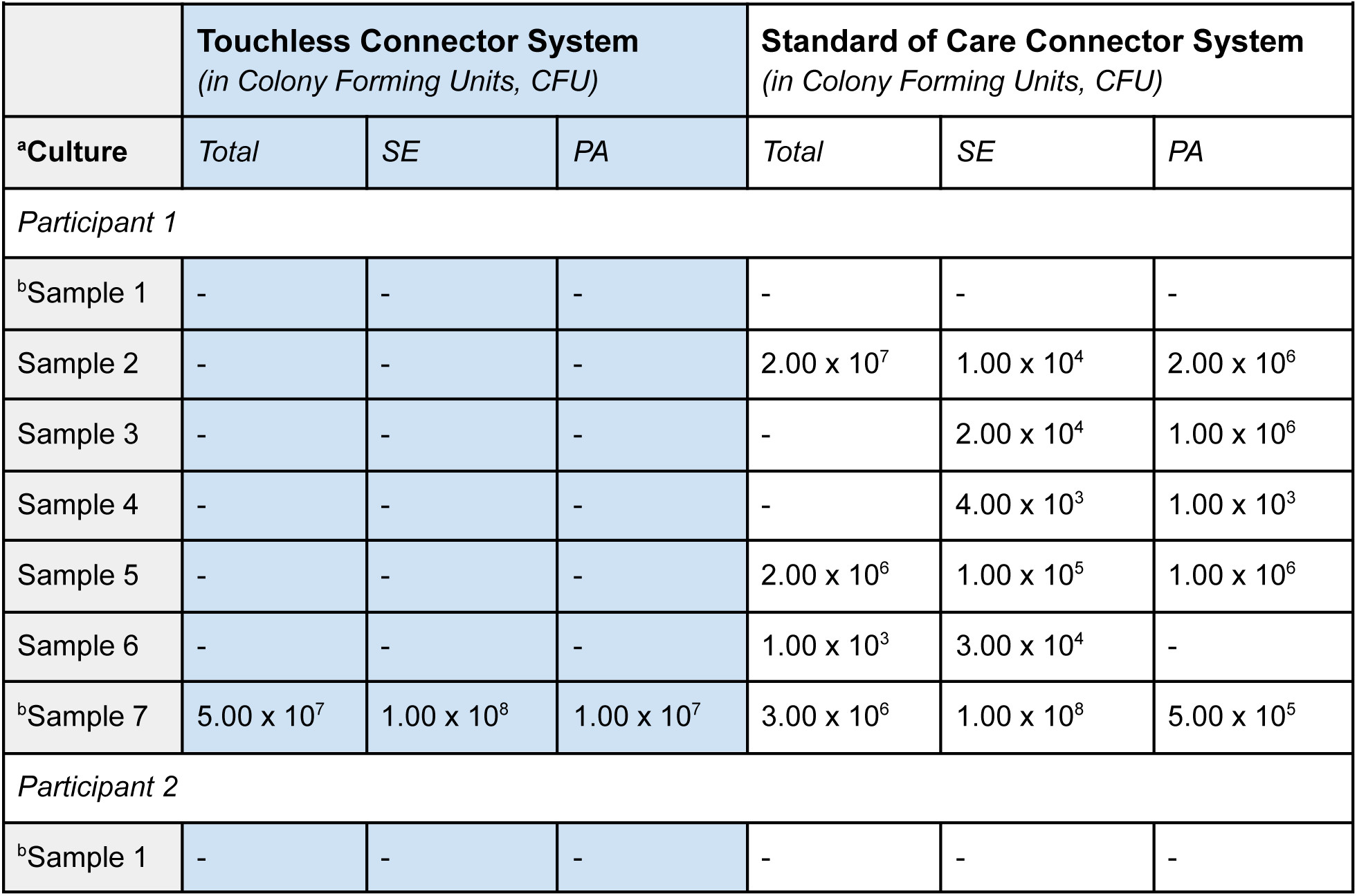
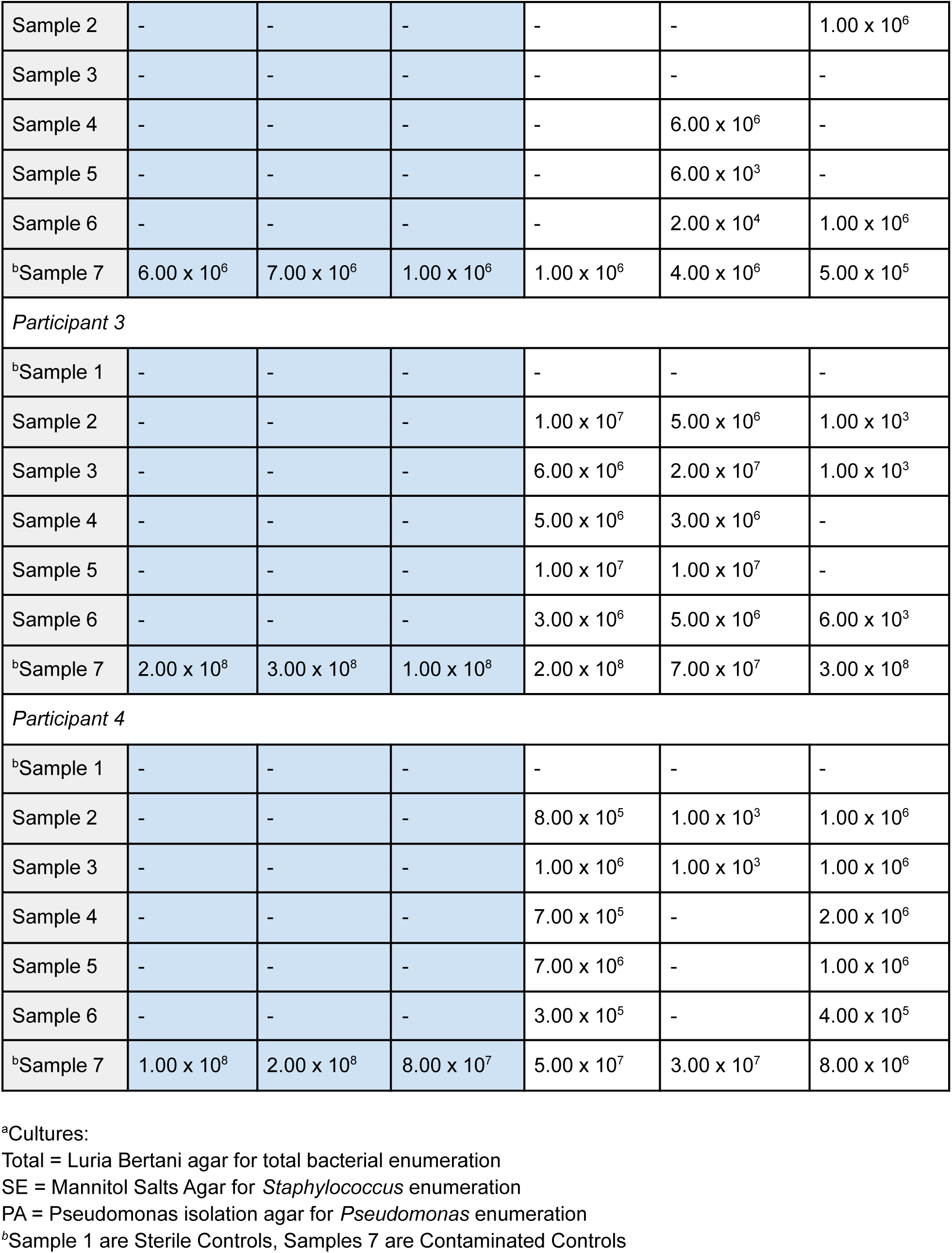
Results of microbiology enumeration for test samples.

| **Table 3: Results of microbiology enumeration for test samples** |  |  |  |  |  |  |
| --- | --- | --- | --- | --- | --- | --- |
|  | **Touchless Connector System**<br>*(in Colony Forming Units, CFU)* |  |  | **Standard of Care Connector System**<br>*(in Colony Forming Units, CFU)* |  |  |
| <sup>a</sup>Culture | *Total* | *SE* | *PA* | *Total* | *SE* | *PA* |
| *Participant 1* |  |  |  |  |  |  |
| <sup>b</sup>Sample 1 | - | - | - | - | - | - |
| Sample 2 | - | - | - | 2.00 x 10<sup>7</sup> | 1.00 x 10<sup>4</sup> | 2.00 x 10<sup>6</sup> |
| Sample 3 | - | - | - | - | 2.00 x 10<sup>4</sup> | 1.00 x 10<sup>6</sup> |
| Sample 4 | - | - | - | - | 4.00 x 10<sup>3</sup> | 1.00 x 10<sup>3</sup> |
| Sample 5 | - | - | - | 2.00 x 10<sup>6</sup> | 1.00 x 10<sup>5</sup> | 1.00 x 10<sup>6</sup> |
| Sample 6 | - | - | - | 1.00 x 10<sup>3</sup> | 3.00 x 10<sup>4</sup> | - |
| <sup>b</sup>Sample 7 | 5.00 x 10<sup>7</sup> | 1.00 x 10<sup>8</sup> | 1.00 x 10<sup>7</sup> | 3.00 x 10<sup>6</sup> | 1.00 x 10<sup>8</sup> | 5.00 x 10<sup>5</sup> |
| *Participant 2* |  |  |  |  |  |  |
| <sup>b</sup>Sample 1 | - | - | - | - | - | - |
| Sample 2 | - | - | - | - | - | $1.00 \times 10^6$ |
| Sample 3 | - | - | - | - | - | - |
| Sample 4 | - | - | - | - | $6.00 \times 10^6$ | - |
| Sample 5 | - | - | - | - | $6.00 \times 10^3$ | - |
| Sample 6 | - | - | - | - | $2.00 \times 10^4$ | $1.00 \times 10^6$ |
| <sup>b</sup>Sample 7 | $6.00 \times 10^6$ | $7.00 \times 10^6$ | $1.00 \times 10^6$ | $1.00 \times 10^6$ | $4.00 \times 10^6$ | $5.00 \times 10^5$ |
| *Participant 3* |  |  |  |  |  |  |
| <sup>b</sup>Sample 1 | - | - | - | - | - | - |
| Sample 2 | - | - | - | $1.00 \times 10^7$ | $5.00 \times 10^6$ | $1.00 \times 10^3$ |
| Sample 3 | - | - | - | $6.00 \times 10^6$ | $2.00 \times 10^7$ | $1.00 \times 10^3$ |
| Sample 4 | - | - | - | $5.00 \times 10^6$ | $3.00 \times 10^6$ | - |
| Sample 5 | - | - | - | $1.00 \times 10^7$ | $1.00 \times 10^7$ | - |
| Sample 6 | - | - | - | $3.00 \times 10^6$ | $5.00 \times 10^6$ | $6.00 \times 10^3$ |
| <sup>b</sup>Sample 7 | $2.00 \times 10^8$ | $3.00 \times 10^8$ | $1.00 \times 10^8$ | $2.00 \times 10^8$ | $7.00 \times 10^7$ | $3.00 \times 10^8$ |
| *Participant 4* |  |  |  |  |  |  |
| <sup>b</sup>Sample 1 | - | - | - | - | - | - |
| Sample 2 | - | - | - | $8.00 \times 10^5$ | $1.00 \times 10^3$ | $1.00 \times 10^6$ |
| Sample 3 | - | - | - | $1.00 \times 10^6$ | $1.00 \times 10^3$ | $1.00 \times 10^6$ |
| Sample 4 | - | - | - | $7.00 \times 10^5$ | - | $2.00 \times 10^6$ |
| Sample 5 | - | - | - | $7.00 \times 10^6$ | - | $1.00 \times 10^6$ |
| Sample 6 | - | - | - | $3.00 \times 10^5$ | - | $4.00 \times 10^5$ |
| <sup>b</sup>Sample 7 | $1.00 \times 10^8$ | $2.00 \times 10^8$ | $8.00 \times 10^7$ | $5.00 \times 10^7$ | $3.00 \times 10^7$ | $8.00 \times 10^6$ |

<sup>a</sup>Cultures:
Total = Luria Bertani agar for total bacterial enumeration
SE = Mannitol Salts Agar for *Staphylococcus* enumeration
PA = Pseudomonas isolation agar for *Pseudomonas* enumeration
<sup>b</sup>Sample 1 are Sterile Controls, Samples 7 are Contaminated Controls

## 4. DISCUSSION

Cultures of samples from the touchless connector system showed no bacterial growth in the relevant connection attempts (i.e. Samples 2 - 6 for all 4 participants).

In comparison, cultures of samples derived from the standard of care showed bacterial growth. Luria Bertani agar for total organisms, showed bacterial growth in 13 of 20 (65%) relevant samples **(Figure 5)**. Mannitol Salts Agar, for *Staphylococcus Epidermidis* showed bacterial growth in 15 of 20 (75%) relevant samples, and Pseudomonas isolation agar for *Pseudomonas aeruginosa* showed bacterial growth in 14 of 20 (70%) relevant samples.

**Figure 5:**
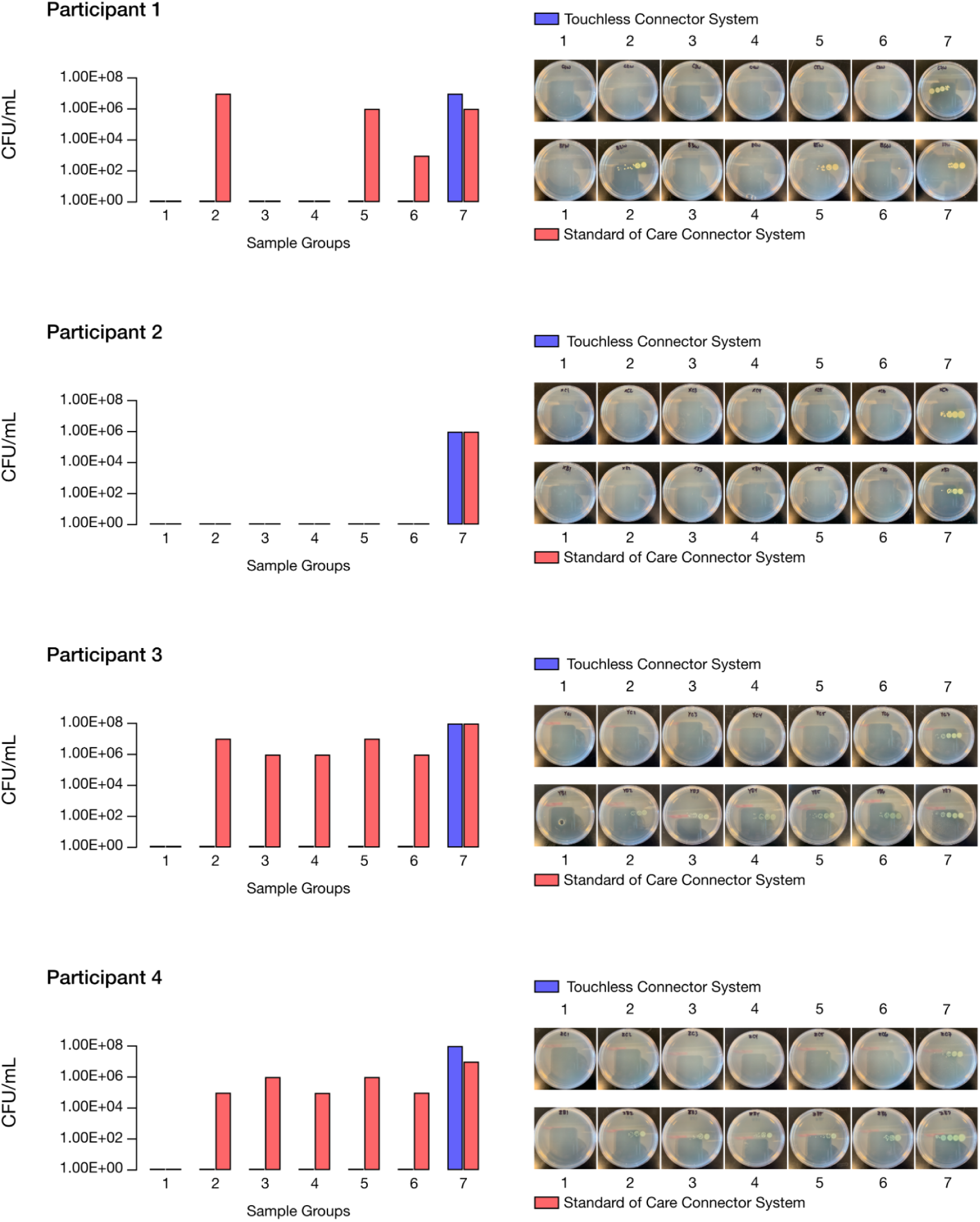
Contamination on Luria Bertani Agar CFU/mL = Colony Forming Units/mL

## 5. CONCLUSION

Results show that the touchless connector system prevents fluid path contamination even with heavy bacterial exposures and may help reduce peritoneal dialysis-associated peritonitis risks from inadvertent contamination with further development.

## Data Availability

Not Applicable.

## Contributions

IOY drafted the work and this was then revised critically by all authors. The experiments were conducted by IOY and TV with the microbiology cultures developed by RH. All authors contributed to the conception and design of the work. All authors had final approval of the version to be published and are jointly accountable for all aspects of the work.

## Acknowledgments

The team would like to recognize the contributions of Amy Hoelscher, Kieran Leong and Yasheen Brijlal to the early concept prototypes. We would also like to thank Heather Collier and the Fairview Kidney Center for providing essential feedback and assistance.

